# Systematic Review and Scientific Rating of Commercial Apps Available in India for Diabetes Prevention

**DOI:** 10.1101/2021.02.15.21251723

**Authors:** H Ranjani, S Nitika, R Hariharan, H Charumeena, N Oliver, R Pradeepa, J C Chambers, R Unnikrishnan, V Mohan, P Avari, RM Anjana

## Abstract

**BACKGROUND:** Scientific evidence for digital health applications (apps) which claim to help in the prevention and management of Type 2 diabetes (T2D) is limited.

**OBJECTIVES:** We aimed to evaluate the quality of currently available health apps for prevention of T2D amongst Asian Indians.

**METHODS:** Using the keywords, ‘diabetes prevention’, ‘healthy lifestyle’ and ‘fitness’, a total of 1486 apps available in India via Google Play were assessed for eligibility by two independent reviewers. After initial screening using specific inclusion and exclusion criteria, 50 apps underwent a pre-specified rating based on user reviews, number of downloads and app size. Sixteen apps that scored ≥ 9 were shortlisted for further review using the Mobile App Rating Scale (MARS). MARS contains 2 main categories: Category I (Application Quality) and Category II (Application Subjective Quality). The mean MARS scores were used to identify the top ranked apps.

**RESULTS:** The mean score for Category I of MARS rating was highest for ‘Google Fit: Health and Activity Tracking’ (4.55/5). This was followed by ‘Healthifyme - Diet Plan, Health and Weight Loss’ (4.45/5). For Category II of MARS, ‘Diabetes M’, ‘Google Fit: Health and Activity Tracking’ and Calorie Counter- My fitness pal’, ‘Healthifyme - Diet plan, Health and Weight Loss’ all scored equally well. On comparing the advantages and disadvantages of each of these applications, ‘Google Fit: Health and Activity Tracking’ and ‘Healthifyme - Diet plan, Health and Weight Loss’ again ranked the best.

**CONCLUSIONS:** Our review identifies two commercially available apps ‘Google Fit: Health and Activity Tracking’ and ‘Healthifyme - Diet plan, Health and Weight Loss’ as being user friendly and good quality. Although encouraging, further research is needed to evaluate the efficacy of these apps for prevention of diabetes.

## Introduction

Mobile technology has changed the life of millions of people around the world [1]. Smart phones provide an ideal combination of user-accessibility with high functionality of customized content. These have already achieved high penetration across Asia. India accounts for 20% of the global smart phone market with approximately 370 million users [2]. It is estimated that the number of smartphone users in India will double to reach between 650 million and 700 million by 2023 [2] rendering mobile health (mHealth) an indispensable component of health care [3].

The term mHealth refers to clinical and public health activities, made available through smartphone devices, which offer health related information and services to people anywhere, anytime [4]. mHealth encourages users to be part of their own health management plan, particularly when it relates to prevention and/or self-management for chronic conditions [5]. The increase in usage of smartphones has led to the rapid development of many mHealth applications. The number of mHealth apps available at Google Play in the first quarter of 2020 was 43285, compared to 23955 in the first quarter of 2015 [6]. These mHealth apps allow people to be connected with their health care providers like never before [7]. International guidelines support the use of mHealth apps for prevention of type 2 diabetes (T2D) and cardiovascular disease (CVD) in high-risk individuals [8,9]. However, little information on the quality of apps is available, and merely selecting health apps on the basis of popularity does not provide any meaningful information on app quality [10,11].

Hence, in this study we aimed to collate data on currently available commercial health apps and evaluate their potential quality as tools for prevention of T2D, with particular reference to Asian Indians.

## Methodology

The systematic review process comprised of three stages:

### Stage 1: Searching the Android platform

Two independent primary reviewers searched for health applications commercially available in India on the android platform (Google Play store) on 3^rd^ June 2020 using three key phrases: ‘diabetes prevention’, ‘healthy lifestyle’, and ‘fitness’. Other key words such as “exercise” and “physical activity” were not included, as these searches yielded apps focusing on exercise routines, strategies related to athletic training and had no link to diabetes prevention. Using the key word “diabetes” alone, yielded apps which helped to diagnose and/or manage diabetes. Thus, we focused primarily on the key words ‘diabetes prevention, healthy lifestyle and fitness’ as they yielded the apps that were relevant to diabetes prevention. In total, 1486 apps in the Google Play store matched with the keywords; 500 apps using the keyword “diabetes prevention”, 498 apps for “healthy lifestyle” and 488 apps for “fitness” *(Figure 1)*. These were assessed for eligibility by the primary reviewers, with each reviewer assessing approximately 750 applications.

**Figure 1:**
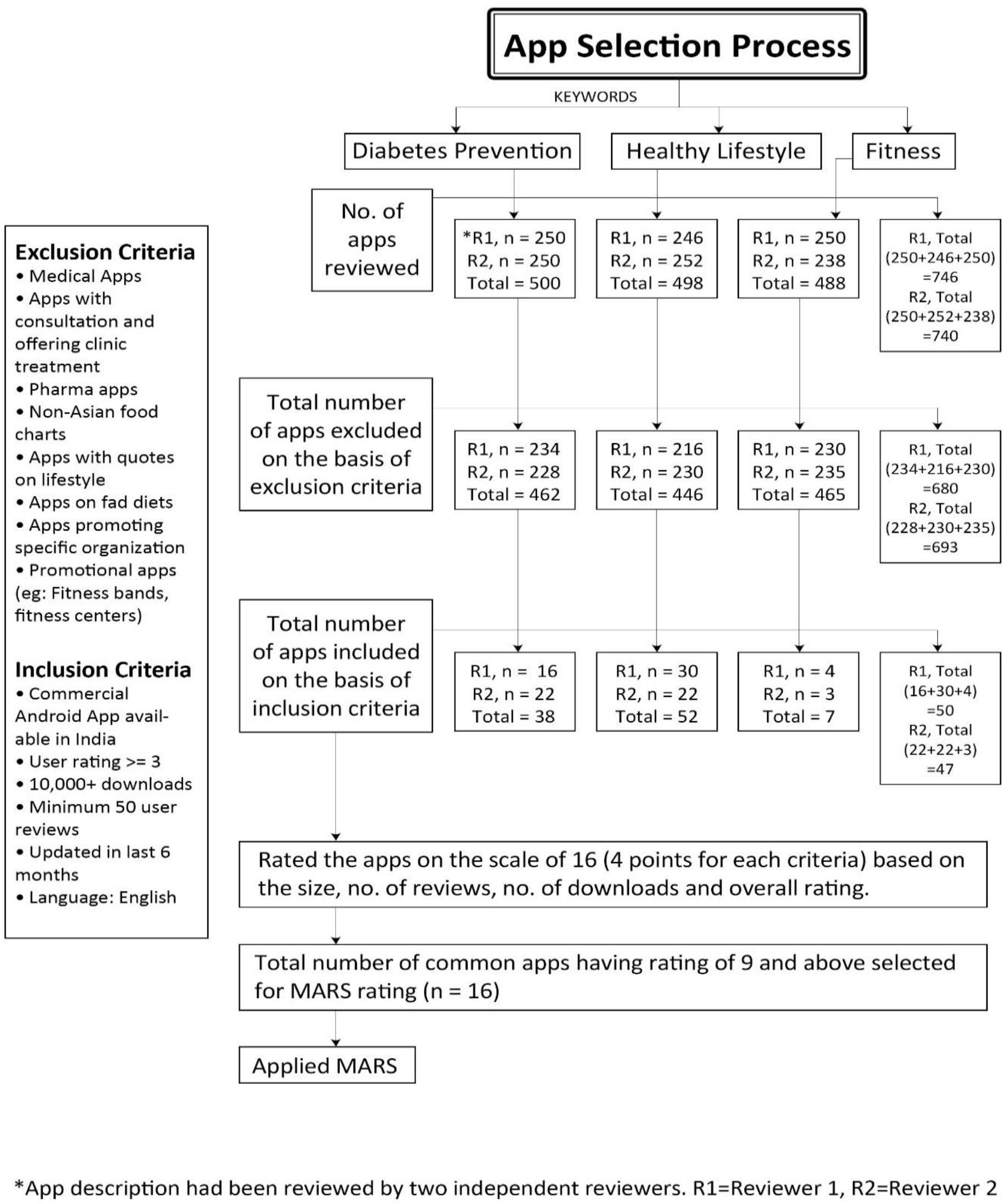
Flow chart to depict the systematic search of the apps on an android platform.

Inclusion criteria were an average user rating ≥ 3, more than 10,000 downloads, minimum 50 user reviews, app updated in the last 6 months and available in the English language. As we were looking for apps which could be used across the country, we preferred to select apps available in English which is the link language across urban India. Exclusion criteria were medical apps providing diagnostic or clinician-led healthcare (for example, ‘DIABNEXT-Make your diabetes management easy’ and ‘Diabetes Diagnostics’), apps with consultation provided (‘Gadge Diabetes Care’), pharmacy apps, non-Asian food charts (apps that focused on American/ western diets and not relevant to Indian cultural preferences or habits), apps with quotes on lifestyle (‘Best Life tips’), apps on unconventional diets for weight loss (‘Total Keto Diet’), apps promoting a specific organization or products (‘Medworks Management of Diabetes’, ‘Lefum Health’, ‘Lenovo life’) and promotional apps (e.g.: Fitness bands, Fitness centres).Premium features requiring payment from the user were excluded, however apps providing a basic version without additional cost were included. Based on these exclusion criteria, reviewer 1 and reviewer 2 excluded 680 and 693 apps respectively. Thus, reviewer 1 shortlisted 50 apps and reviewer 2, 47 apps *(Figure 1)*.

Subsequently, detailed reviews posted by users were explored to study the app features and specifications. A rating scale (*Appendix 1*) was applied based on the following 4 parameters that reflect their availability in the Play store: - a) overall rating of the app, b) number of downloads, c) size of the app and d) number of reviews. Each primary reviewer rated their list of apps on a scale of 1 to 4 [1 being the lowest and 4 being the highest, total rating out of 16 (4*4)]. There were16 apps that received a total rating score ≥9 from both reviewers, and these were shortlisted for further evaluation (*Appendix 2*).

### Stage 2: Call for commercial apps

Developers of commercial health apps (apps which already have been published in the Google Play store) were invited to participate in our Digital Health Intervention study. This digital intervention study is the next stage of the project, wherein the two commercial mHealth apps selected through this systematic review process and two in-house apps will be tested for usability, preferences, engagement, and weight loss in 1000 participants. This was done “simultaneously” along with stage 1. There could have been relevant apps which were missed in our search due to the descriptions given in the play store. Hence, an official invite to participate was sent to the developers. An advertisement was published on our official website and also on social platforms, including Facebook, Instagram and LinkedIn. We received 106 responses but only one response lead was relevant to our study (‘Fittr’). To increase the number of responses, we did another process of Search Engine Optimization. This process added 20 more apps, out of which 11 had already been shortlisted by the two primary reviewers in their initial search. The remaining 9 apps were reviewed and rated (as performed in Stage 1) but none scored above 9.

A secondary reviewer reviewed the apps shortlisted by the two primary reviewers to verify and solve any disputes regarding the final 16 apps selected from stages 1 and 2. Stages 1 and 2 were performed simultaneously.

### Stage 3: Applying MARS

The Mobile App Rating scale (MARS) is a medical app quality rating tool that provides a multidimensional assessment of app quality indicators like engagement, functionality, aesthetics, and information quality, as well as app subjective quality (*Figure 2*) [11,12]. MARS contains 2 main categories. The developers of this scale have not divided MARS into categories but have mentioned divisions such as ‘Application Quality’ (Sections A to D) and ‘Application Subjective Quality’ (Section E). For ease of understanding, we have renamed ‘Application Quality’ as Category I and ‘Application Subjective Quality as Category II’. Category I assesses quality across various dimensions. It contains 4 sections (A-D) and all items are rated on a 5-point scale from “1. Inadequate” to “5. Excellent”. A mean score is given at the end of each section (A-D). These mean scores are averaged to get an overall mean score for these categories (out of 5) as the app quality mean score.

**Figure 2:**
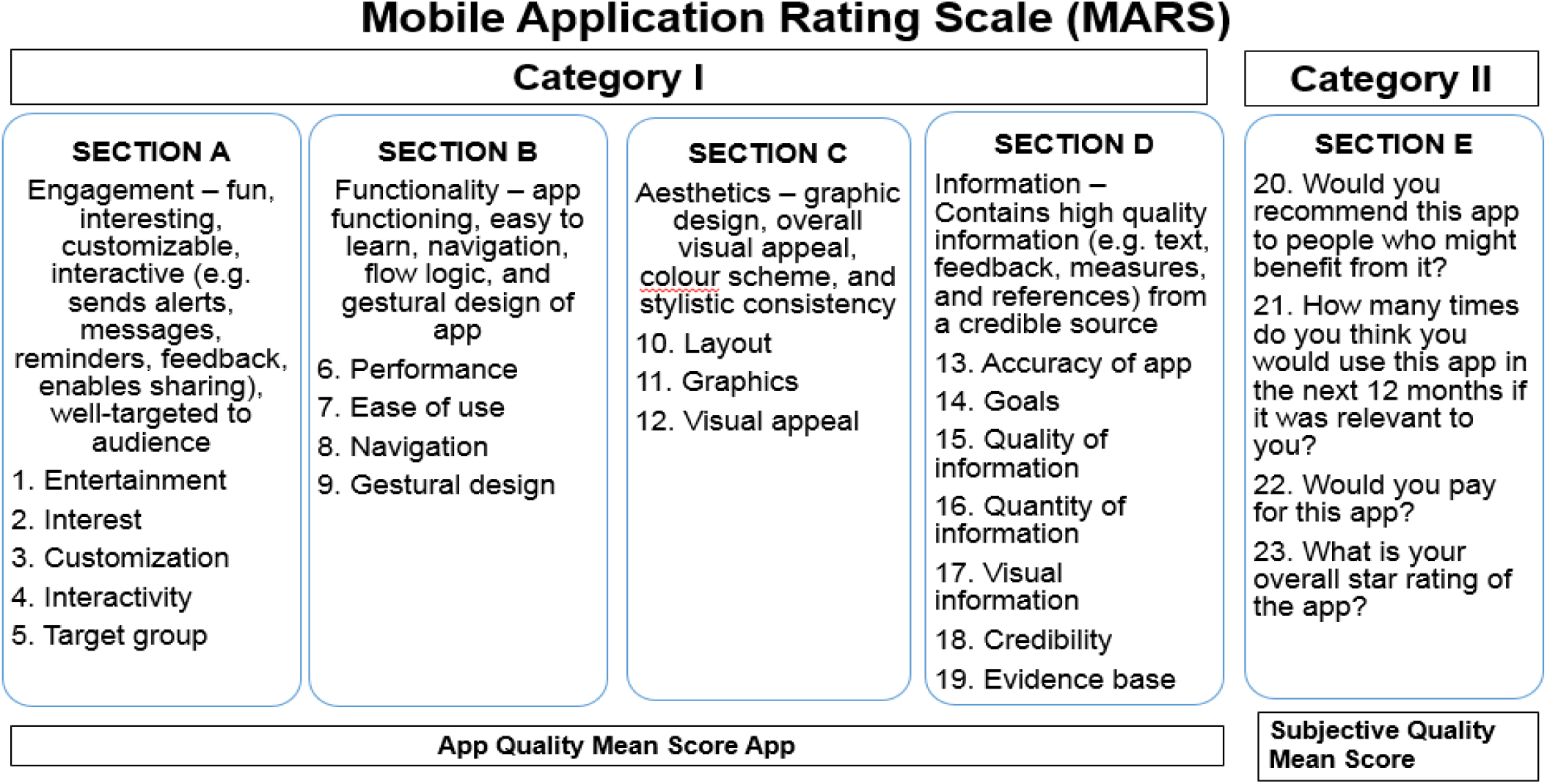
Categories of MARS.

Category II is the Application Subjective Quality that has questions (Section E) on recommendation, frequency of use, willingness to buy the app and the overall rating given for the app from the user point of view. The apps are scored and presented separately category wise.

The two primary reviewers independently downloaded the 15 shortlisted apps (1 app could not be downloaded due to technical issues) on their android phones and used them extensively to conduct in-depth evaluations of each app before rating them using MARS. The reviewers were trained to use the MARS instrument by watching an online tutorial [13], to ensure rating consistency amongst the reviewers. The mean MARS scores were used to identify the best quality apps. Kappa statistic was used to estimate inter-reviewer (rater) reliability between the two primary reviewers [14]. The reliability estimate was 0.77 (*Cohen Kappa calculated using the SPSS version 15*). As the domains in this scale are subjective in nature, this reliability estimate was considered to be very good.

## Results

The 15 apps which were downloaded are listed below:

1. Google Fit: Health and Activity Tracking
2. Healthifyme - Diet Plan, Health and Weight Loss
3. Calorie Counter - My fitness pal
4. Habits Diabetes coach
5. Diabetic Recipes: Healthy Food
6. Calorie Counter - MyNetDiary, Food diary tracker
7. 8fit Workouts and Meal Planner
8. Beat O SMART Diabetes Management
9. Diabetic Diet Recipes : Control Diabetes and Sugar
10. Diabetes:M
11. Noom: Health and Weight
12. mySugr-Diabetes App and Blood Sugar Tracker
13. Diabetes Forum
14. Diabetes Diary - Blood Glucose Tracker
15. Beat Diabetes

*Figure 3* shows the results of MARS for App Quality (sections A-D). In section A, Engagement, ‘Google Fit: Health and Activity Tracking’ had the highest engagement mean score of 4.50/5 for including the notification features for activity tips, goal progress tips, goals adjustment and completed goals. Meanwhile, ‘Beat Diabetes’ had the lowest *Engagement mean score* of 2.30/5. This app had limited information and was less interactive.

**Figure 3:**
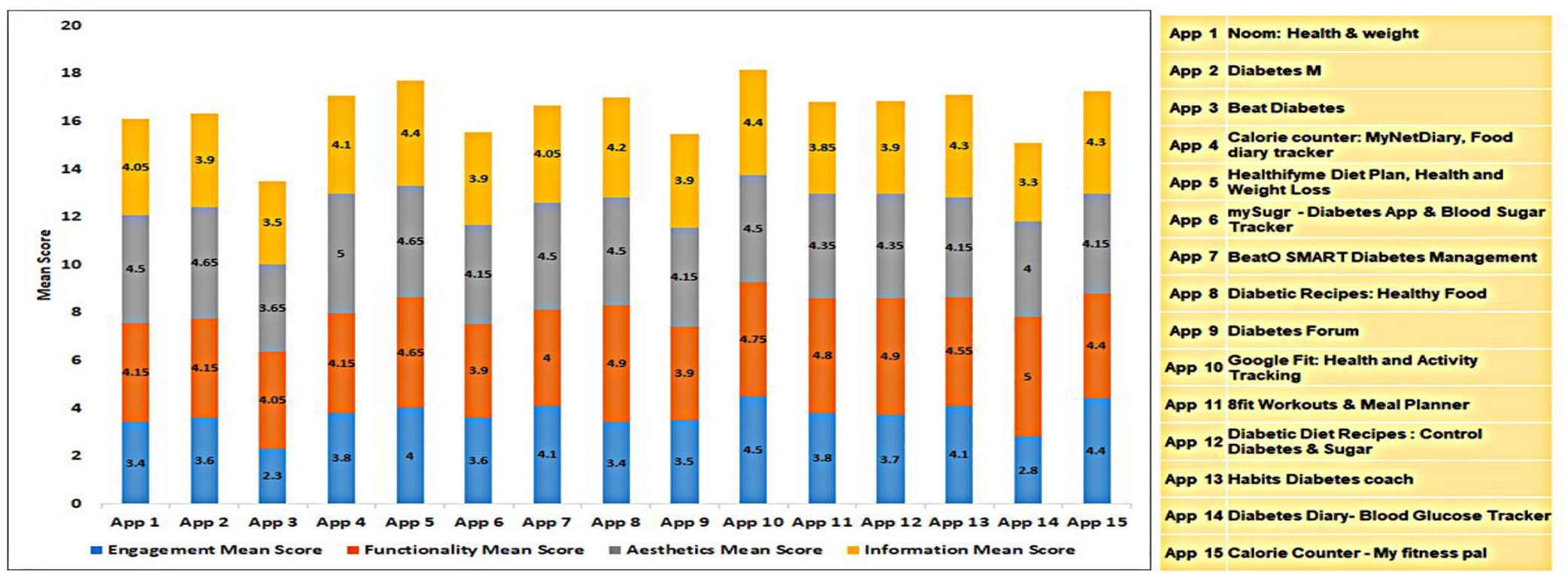
Mean score of MARS Sections A- App Engagement, B- Functionality, C- Aesthetics and D- Information (all 15 apps)

In section B; Functionality, ‘Diabetes Diary: Blood Glucose Tracker’ scored the highest on functionality, with a mean score of 5.00/5. ‘mySugr: Diabetes App and Blood Sugar Tracking’ and ‘Diabetes Forum’ had the lowest *Functionality mean score* of 3.90/5. ‘mySugr: Diabetes App and Blood Sugar Tracking’ required an external glucometer to be connected and ‘Diabetes Forum’ had glitches in the registration and user log in processes.

For section C; Aesthetics, the mean score for all the apps was above 4.00. ‘Calorie Counter: MyNetDiary, Food diary tracker’ had the highest score of 5.00/5. The app had a clear and orderly organized layout with high quality graphics and visual design. The colour scheme further enhanced the app features. On the other hand, ‘Beat Diabetes’ had the lowest *Aesthetics mean score* of 3.65/5.

In section D; Information, ‘Healthifyme’ and ‘Google Fit: Health and Activity Tracking’ had the highest score of 4.40/5. Both the apps provided a thorough description of the app components and had specific and achievable goals. The quality of information was well written and relevant to the goals of the app. Visual information was clear and logical. ‘Google Fit: Health and Activity Tracking’ App also was evidence-based [15]. The ‘Diabetes Diary: Blood Glucose Tracker’ app had the lowest *Information mean score* of 3.30/5. This app did not appear to have specific, measurable goals.

The mean MARS scores for each of the 15 apps downloaded by the two primary reviewers are shown in *Table 1*. According to the mean scores of both reviewers for Category I, ‘Google Fit: Health and Activity Tracking’ was ranked at number 1 with the average app quality mean score of 4.55. This was followed by ‘Healthifyme - Diet Plan, Health and Weight Loss’ with a mean score of 4.45. Two apps, namely ‘Calorie Counter - My fitness pal’ and ‘Habits Diabetes coach’ were tied at number 3 (mean score 4.30). These were followed by ‘Calorie Counter - MyNetDiary, Food Diary Tracker’ and ‘Diabetic Recipes: Healthy Food’ (mean score 4.25) at number 4, and ‘Diabetic Diet Recipes: Control Diabetes and Sugar’, ‘Beat O SMART Diabetes Management’ and ‘8fit Workouts and Meal Planner’ (mean score 4.20) at number 5.

**Table 1:**
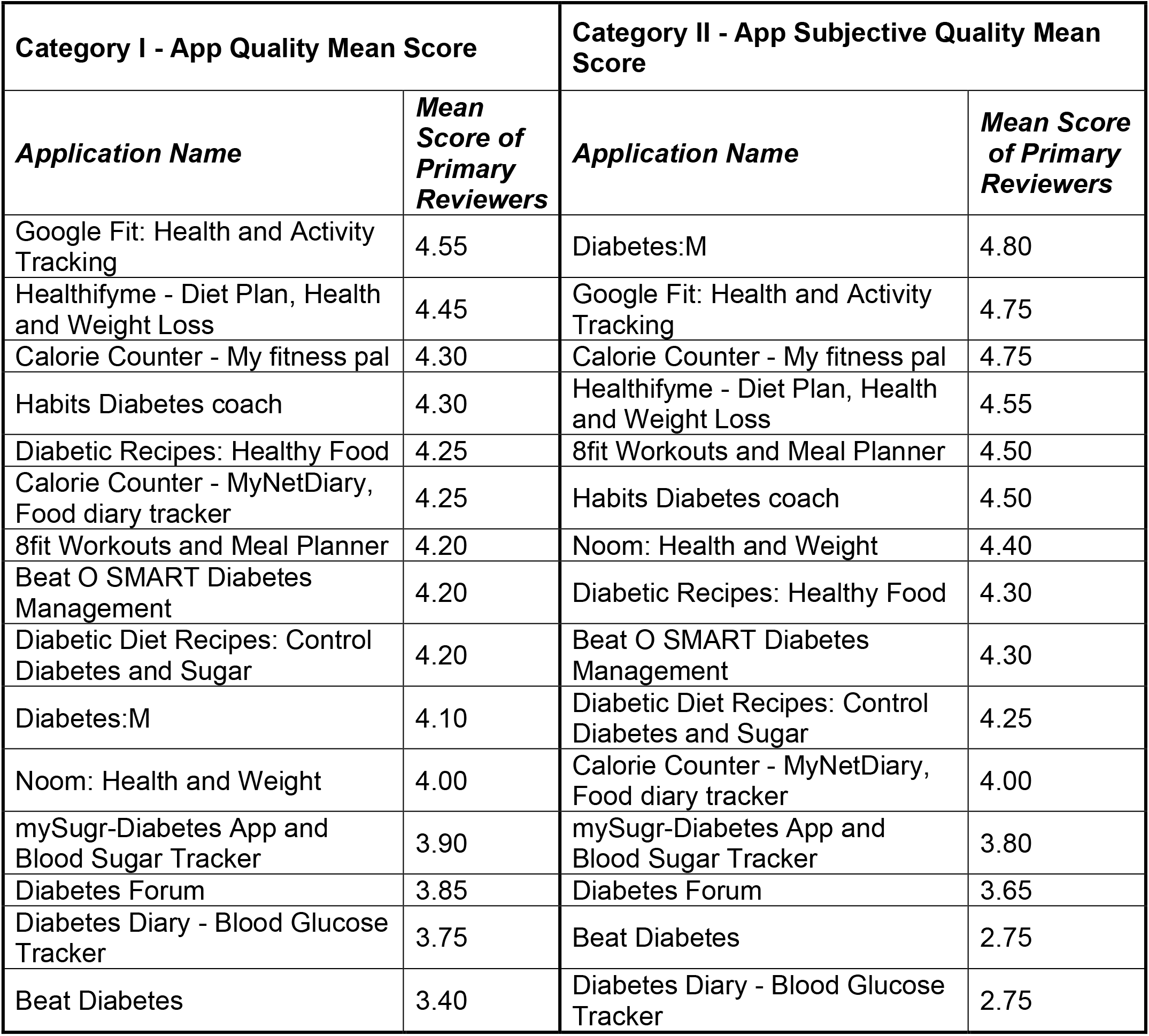
Overall mean MARS scores under each category for the 15 apps.

| **Category I - App Quality Mean Score** |  | **Category II - App Subjective Quality Mean Score** |  |
| --- | --- | --- | --- |
| ***Application Name*** | ***Mean Score of Primary Reviewers*** | ***Application Name*** | ***Mean Score of Primary Reviewers*** |
| Google Fit: Health and Activity Tracking | 4.55 | Diabetes:M | 4.80 |
| Healthifyme - Diet Plan, Health and Weight Loss | 4.45 | Google Fit: Health and Activity Tracking | 4.75 |
| Calorie Counter - My fitness pal | 4.30 | Calorie Counter - My fitness pal | 4.75 |
| Habits Diabetes coach | 4.30 | Healthifyme - Diet Plan, Health and Weight Loss | 4.55 |
| Diabetic Recipes: Healthy Food | 4.25 | 8fit Workouts and Meal Planner | 4.50 |
| Calorie Counter - MyNetDiary, Food diary tracker | 4.25 | Habits Diabetes coach | 4.50 |
| 8fit Workouts and Meal Planner | 4.20 | Noom: Health and Weight | 4.40 |
| Beat O SMART Diabetes Management | 4.20 | Diabetic Recipes: Healthy Food | 4.30 |
| Diabetic Diet Recipes: Control Diabetes and Sugar | 4.20 | Beat O SMART Diabetes Management | 4.30 |
| Diabetes:M | 4.10 | Diabetic Diet Recipes: Control Diabetes and Sugar | 4.25 |
| Noom: Health and Weight | 4.00 | Calorie Counter - MyNetDiary, Food diary tracker | 4.00 |
| mySugr-Diabetes App and Blood Sugar Tracker | 3.90 | mySugr-Diabetes App and Blood Sugar Tracker | 3.80 |
| Diabetes Forum | 3.85 | Diabetes Forum | 3.65 |
| Diabetes Diary - Blood Glucose Tracker | 3.75 | Beat Diabetes | 2.75 |
| Beat Diabetes | 3.40 | Diabetes Diary - Blood Glucose Tracker | 2.75 |

From Category II of the MARS ‘Diabetes M’ had the highest App Subjective Quality Mean Score of 4.80/5, scoring highly on all the parameters, including app recommendation, frequency of use, willingness to buy and the overall rating. Conversely, the two apps ‘Diabetes Diary: Blood Glucose Tracker’ and ‘Beat Diabetes’ had the lowest App Subjective Quality Mean Score of 2.75/5. ‘Beat Diabetes’ scored lowest for willingness to buy. Thus, ‘Diabetes M’, ‘Google Fit: Health and Activity Tracking’ and Calorie Counter-My Fitness Pal’ and ‘Healthifyme’ were the top ranked apps as per their App subjective quality mean scores. This was followed by ‘8fit Workouts and Meal Planner’, and ‘Habits Diabetes coach’ tied at number 4 and ‘Noom: Health and Weight’ at number 5. However, an in-depth evaluation of the apps revealed that ‘Diabetes M’ may not be suitable for the Indian cuisine (needs cultural adaptation) and was difficult to access. ‘Healthifyme’ had more features (tracked weight, activity, food and water and provides a personal coach) when compared to ‘Calorie Counter-My Fitness Pal’. Hence, our top ranked apps considering both the ‘App quality mean score’ (Category I) and ‘App subjective quality mean score’ (Category II) were Google fit and Heathifyme.

## Discussion

To our knowledge, this is the first study, to conduct a systematic review and compare quality of mHealth apps available in the Play Store for diabetes prevention in Asian Indians, using a standardized rating tool. We found two commercially available apps ‘Google Fit: Health and Activity Tracking’ and ‘Healthifyme - Diet plan, Health and Weight Loss’ ranked highly amongst the apps available in the Google play store for prevention of diabetes in Asian Indians.

Lifestyle factors such as diet, physical inactivity, excess alcohol consumption, cigarette smoking, drug abuse, stress affecting mental health and lack of sleep are widely recognized as major determinants of metabolic diseases including obesity, T2D and CVD in most populations [16, 17]. Most of these lifestyle behaviours form the base for developing an mHealth app [7]. After analysing the 15 shortlisted apps with respect to these health behaviours, we found that majority dealt with diet and/or physical activity and alluded very briefly to the topics of mental health, stress, sleep and other lifestyle behaviours.

The ‘Google Fit: Health and Activity Tracking’ app ranked well because although it is focussed on physical activity, it scored highly on *Engagement* and *Functionality*. The app is intuitive to use and records physical fitness activities, such as walking, running and cycling. It uses this information to estimate calories burned. Other features offered by the app include weight history, duration of exercise, heart rate monitor and sleep tracker (with the help of a third-party applications), as well as enabling personalized goal settings, with customized tips and actionable coaching. Reports on the Play store suggest there were over five million downloads within 6 months of its release. ‘Google Fit: Health and Activity Tracking’ has previously been reported to be an “effortless and affordable activity tracker with the potential to significantly extend the number of activity tracker users compared to other devices” [15]. The main disadvantage of this app was that it does not track food, however it can connect to many other apps and devices providing a common platform to bring data from various sources together.

Next were ‘Healthifyme and Calorie Counter - My fitness pal’. ‘Healthifyme’ ranked better than ‘Calorie Counter-My Fitness Pal’ in its *Functionality* and *Information*. ‘Healthifyme’ focuses on weight loss, fitness and diabetes prevention through behavioural lifestyle changes. The app sends the user reminders to track their weight, activity, food and water to meet fitness goals. The app also provides personalized reminders for walking and workout and has an artificial intelligence (AI) powered smart nutritionist. The premium version of the app offers personalized coaching by diet, fitness and yoga experts. The app also has success stories to motivate users, as well as more than 500 recipes and nutritional information. In a recent publication [18] by an economist reviewing information technology for primary healthcare in India, ‘Healthifyme’ has been called a promising behavioural application; however, research backed evidence of its impact is not yet available.

Other apps that scored highly were ‘Habits Diabetes coach’, ‘8fit Workouts and Meal Planner’, ‘Noom: Health and Weight’, ‘Diabetes:M’ and ‘Calorie Counter – MyNetDiary, Food Diary Tracker’. ‘Habits Diabetes coach’ can track glucose, activity, diet and weight, give medication reminders, has videos and a lifestyle coach for better engagement. ‘8fit Workouts and Meal Planner’ has an attractive user interface, good quality video content and categorized workouts, including yoga, strength and high intensity exercise. On the other hand, ‘Noom: Health and Weight’ has an in-built steps counter, food log, graphs to track weight and customized course plans. ‘Diabetes:M’ has lot of features with detailed report generation, which include weight, pulse, blood pressure, cholesterol, Hb1Ac and sites of injection. This app allows the user to maintain multiple profiles, has a smart assistant, advises on insulin bolus dose, can add reminders, give options for data management and data sharing, discussion groups and track blood glucose levels. ‘Calorie Counter –MyNetDiary, Food Diary Tracker’ has meals and activity log, barcode reader, weight chart, basic weight loss plan, vitamin and pulse rate log, daily calorie budget and feature to upload recipes. However, there are limited workout plans, complicated graphs, charts and food tracker, lack of Indian food data and limited features in the basic version of the app affected overall *App Quality, Engagement and Information scores* of these apps.

All the apps reviewed in this study had a mean MARS score above 3.75 except for one. This suggests the apps had an overall acceptable level of quality. On evaluating the individual aspects of the app quality mean score, we found the mean score for *Engagement* to be the lowest (3.6) for all the 15 apps; followed by *Information* (4.0). The mean score for *Aesthetics and Functionality* was much higher at 4.4. This indicates that app engagement and information provided by the app are potential target areas for improvement. Similar outcomes were reported in another systematic review on mindfulness apps by Mani et al, where mean engagement scores were low, with aesthetics and information scales found to be moderate [19]. A review of commercial apps available for weight loss, also showed that *Information* scored the lowest amongst the MARS domains, indicating an overall lack of evidence-based content [20]. This suggests further investment by developers in evidence-based, data-driven content, combined with change techniques known to be effective in changing relevant behaviour patterns, is required. This may improve the overall app quality, regardless of the perceived aesthetic qualities of the app.

Published literature has shown that most apps available in the market more or less resemble each other in features [21]. But most of these apps lack authentic information on diabetes and/or obesity prevention, the link between lifestyle behaviours and metabolic disease prevention, as well as suffer from lack of culturally relevant tips and suggestions. A recent mHealth study [22] reported that personalization seems to be the key element in engaging a diverse group of participants. In another study [23], the authors concluded that user experience plays an important role in improving their engagement with the app. Furthermore, user interfaces with real-time data interpretation and smart algorithms may be useful in enhancing user experience, thereby improving engagement. The need to develop new mhealth apps arise from the lacunae that exists with regards to lack of scientific evidence, measurable quality standards and a value proposition for existing apps [24,25]. While we were able to identify two high-quality commercial apps, there is, at present, no evidence available for their efficacy in prevention of T2D. Hence, it is recommended that app developers follow a step wise approach to developing any app: define the problem and end user, (2) review the literature extensively, (3) transform information to knowledge, (4) have a proper security and privacy policy, and (5) evaluate usability and efficacy through scientific means [26, 27]. Another important recommendation to mhealth app developers is that while culturally/linguistically adapting an app, it would be best to have a central base application in which app users could just select language etc. and use. Creating multiple applications to handle language and cultural variations could lead to several development and maintenance issues.

Our study has some limitations. The search terms used in the study were restricted to mhealth apps commercially available in India. Also, the search conducted in Google Play store (for Android phones) might have restricted the results of this review as some of the apps are only found in the App Store (for Apple phones). This study evaluated the basic version available of the selected apps, which would be widely accessible to general population. However, we acknowledge the limitation that paid premium versions may be available for download. In addition, considering the dynamic development of apps, popularity and ratings may change very quickly over time.

## Conclusions

Whilst an increasing number of lifestyle and behaviour change apps are being developed for diabetes prevention and management, the scientific evidence base underlying their use is still limited. Our review identifies two commercially available apps ‘Google Fit: Health and Activity Tracking’ and ‘Healthifyme - Diet plan, Health and Weight Loss’ as being highly rated for their usability. Further research through randomised controlled trials is required to evaluate the usability of these apps in diabetes prevention in Asian Indians.

## Data Availability

I take full responsibility for this data.

## Author Contributions

Conceptualization of paper and writing the initial draft: HR and RMA; Methodology: SN, RH, HC, HR; Data Curation: RH, SN, RMA, PA; Review & Editing: All authors. All authors have read and approved the final version of the manuscript, and agree with the order of presentation of the authors.

## Acknowledgments

At Imperial College London, the infrastructure support was provided by the NIHR Imperial Biomedical Research Centre and the NIHR Imperial Clinical Research Facility. The views expressed are those of the author(s) and not necessarily those of the NHS, the NIHR or the Department of Health and Social Care.

## Abbreviations

AI: artificial intelligence
Apps: applications
CVD: cardiovascular disease
MARS: Mobile App Rating Scale
mHealth: mobile health
T2D: type 2 diabetes

## Appendix 1: Initial rating/scoring scale developed internally based on available information in the google play store

| **Initial Rating Scale** |  |
| --- | --- |
| **Overall app rating in the play store** | **Score out of 4** |
| ≤3.4 | 1 |
| 3.5-3.9 | 2 |
| 4-4.4 | 3 |
| 4.5-5 | 4 |
| **No. of Downloads** |  |
| 10,000-49,999 | 1 |
| 50,000-99,999 | 2 |
| 1,00,000-9,99,999 | 3 |
| ≥10,00,000 | 4 |
| **Size of the app** |  |
| >75MB | 1 |
| 51-75MB | 2 |
| 26-50MB | 3 |
| ≤25MB | 4 |
| **User Reviews (shows popularity)** |  |
| ≤1000 | 1 |
| 1001-4999 | 2 |
| 5000-13999 | 3 |
| ≥14000 | 4 |

## Appendix 2: Basic description and review of the final 15 apps using the play store data (or Initial rating scale given in Appendix 1)

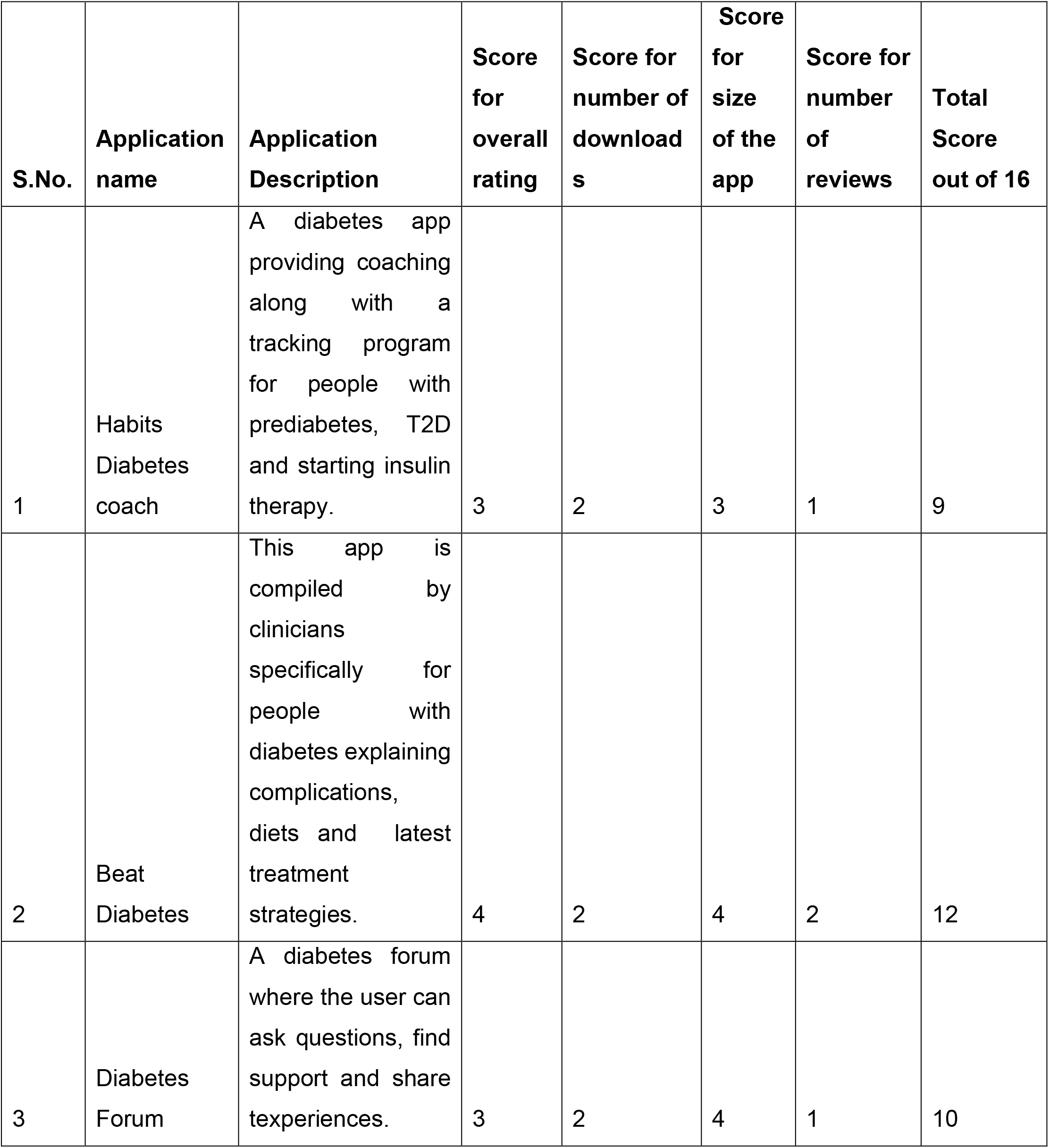
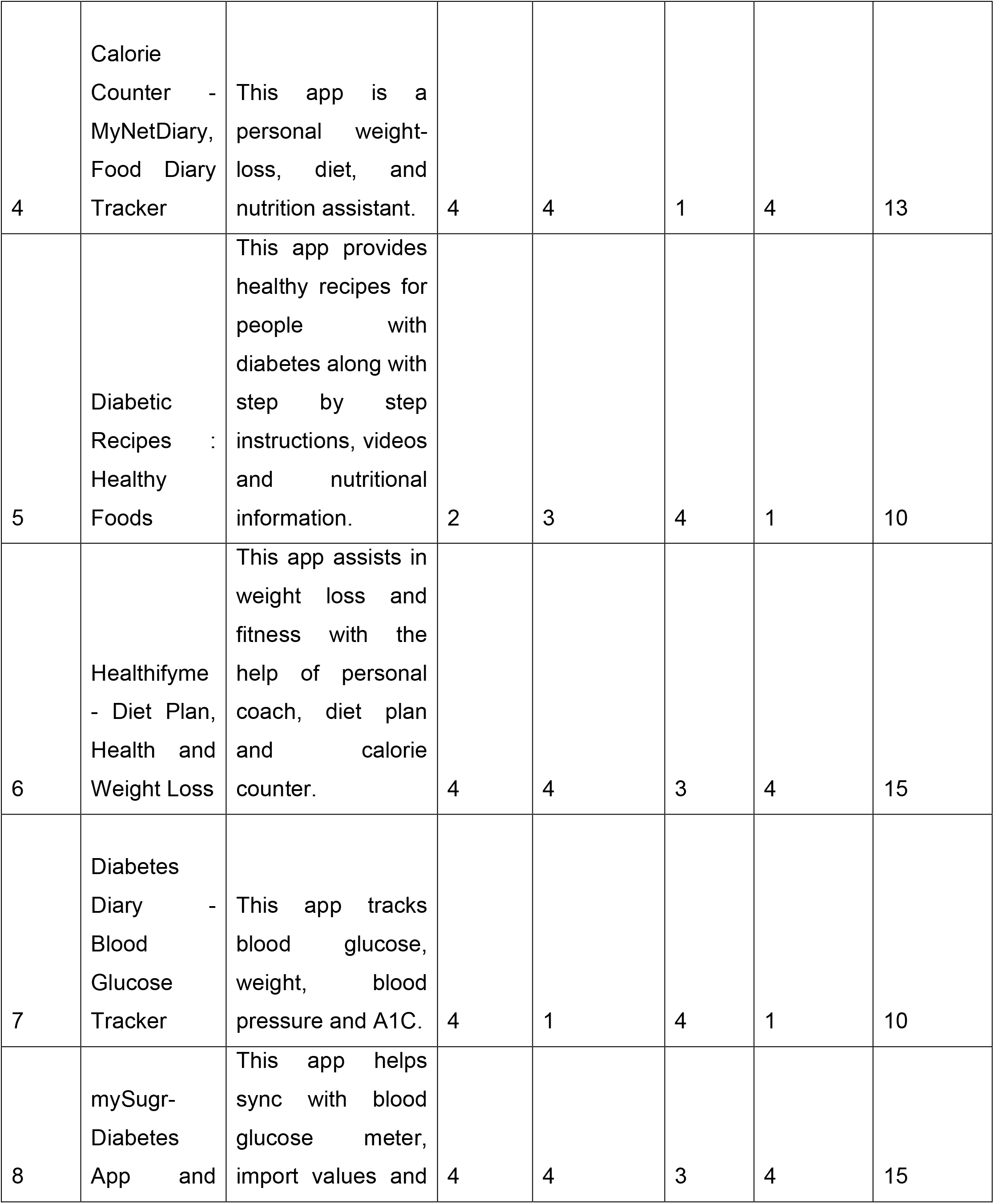
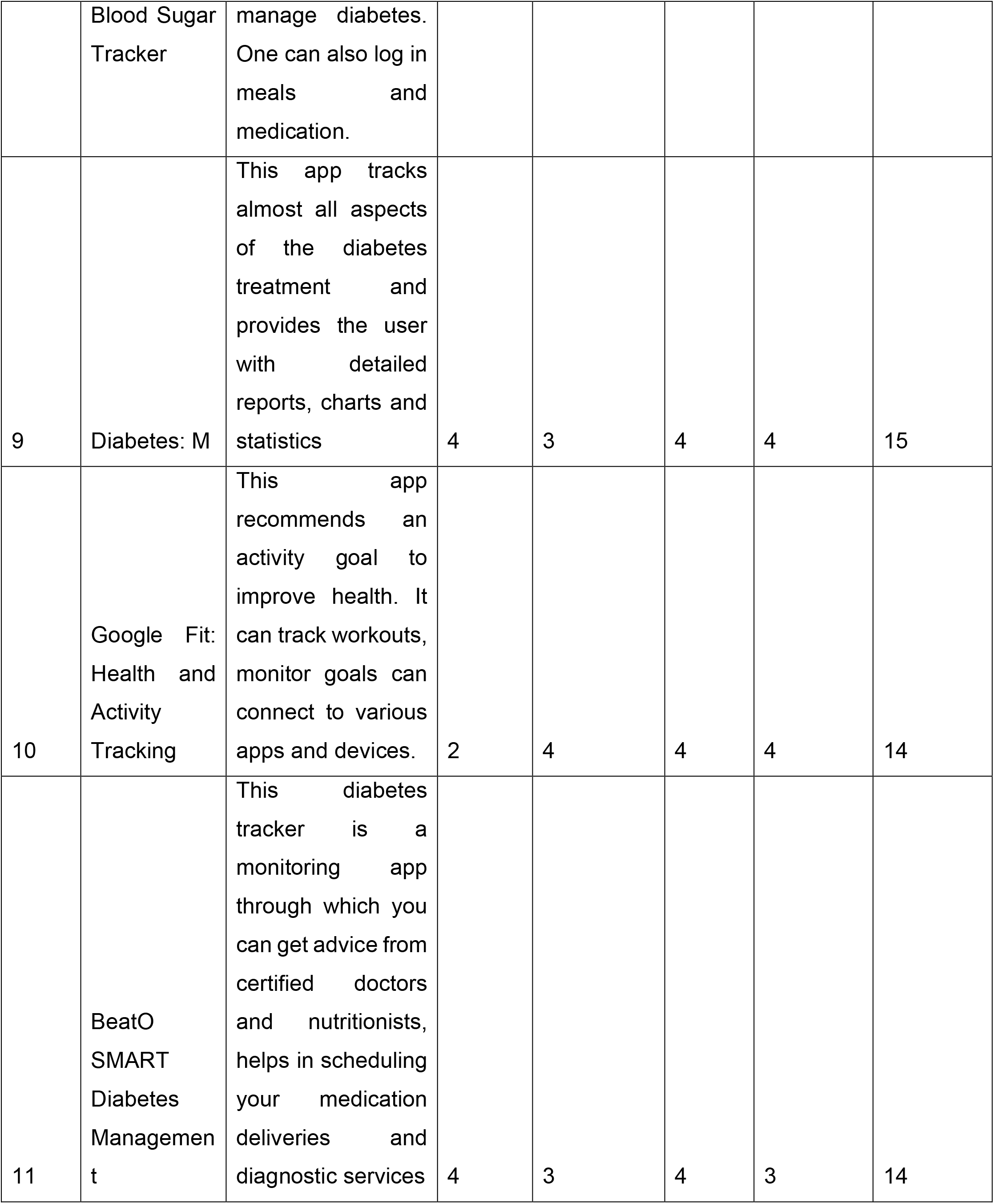
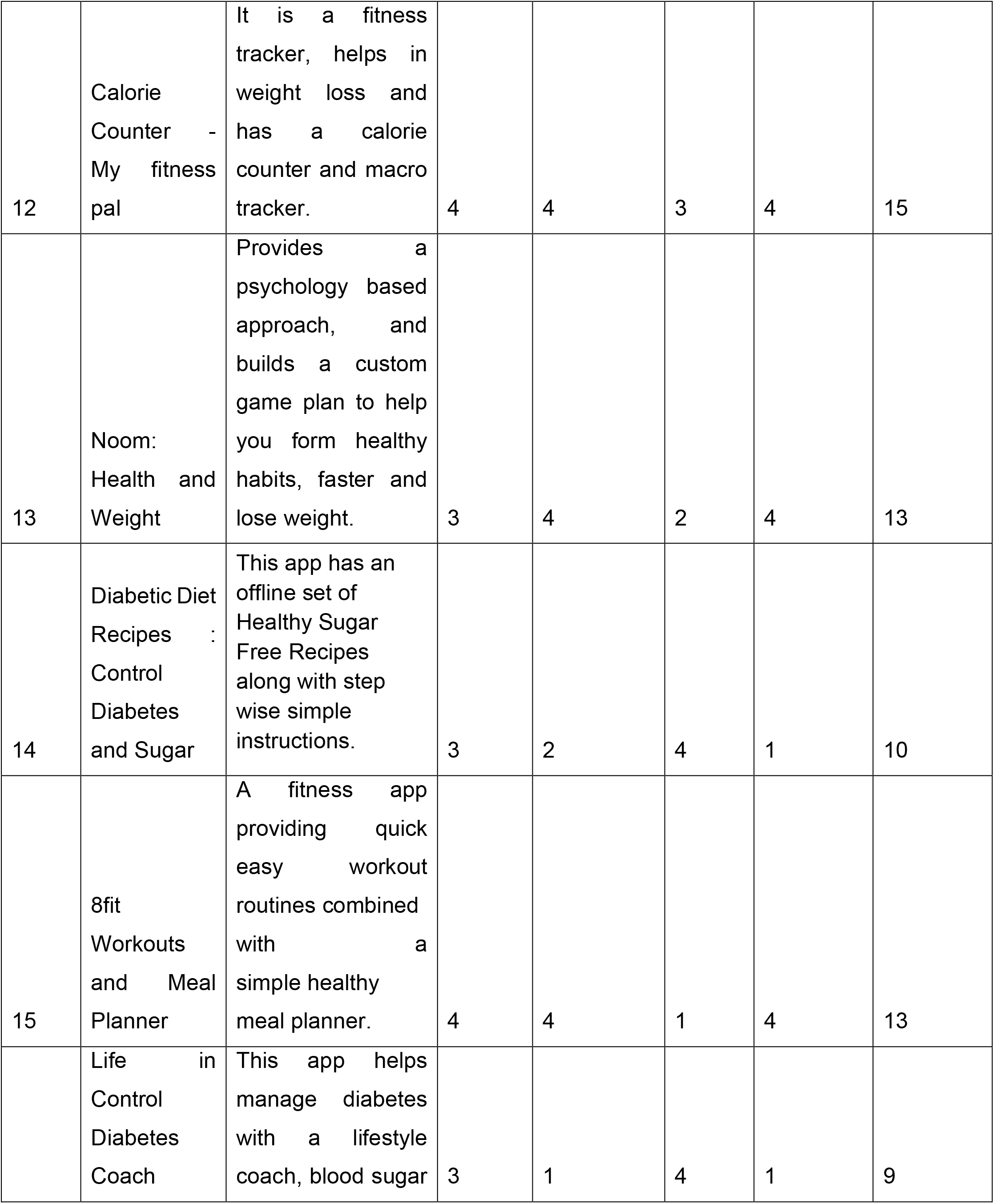
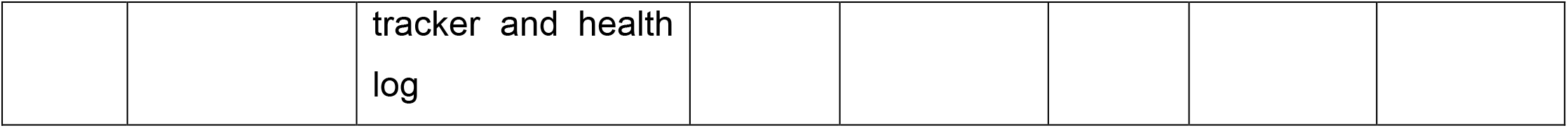

## References

1. Jusoh, S. A survey on trend, opportunities and challenges of mHealth apps. International Journal of Interactive Mobile Technologies (iJIM). 2017; 11: 73–85.

2. IAMAI & KPMG Report “India On The Go – Mobile Internet Vision Report 2017”. URL:http://eeind.in/mobile-internet-use-in-india-2015-16/. Published 2015. Assessed September 22, 2020.

3. Farrington C, Aristidou A, Ruggeri K. mHealth and global mental health: still waiting for the mH2 wedding?.Global Health. 2014;10:17.

4. Klasnja P, Pratt W. Managing health with mobile technology. Interactions. 2014; 21 (1). DOI:10.1145/2540992.

5. Sittig S, Wang J, Iyengar S, Myneni S, Franklin A. Incorporating Behavioral Trigger Messages Into a Mobile Health App for Chronic Disease Management: Randomized Clinical Feasibility Trial in Diabetes. JMIR mHealth and uHealth. 2020; 8: e15927.

6. Matej Mikulic. Google Play: number of available medical apps as of Q4 2020.Published May 13, 2020. URL:https://www.statista.com/statistics/779919/health-apps-available-google-play-worldwide/. Assessed June 26, 2020.

7. Milne-Ives M, Lam C, De Cock C, Van Velthoven MH, Meinert E. Mobile Apps for Health Behavior Change in Physical Activity, Diet, Drug and Alcohol Use, and Mental Health: Systematic Review. JMIR MhealthUhealth. 2020;8(3):e17046.

8. WHO. Global action plan for the prevention and control of NCDs 2013-2020. http://www.who.int/nmh/publications/ncd-action-plan/en/. Assessed August 8, 2020.

9. Cummings E, Borycki E, Roehrer E. Issues and considerations for healthcare consumers using mobile applications. Stud Health Technol Inform. 2013;183:227– 231.

10. Girardello A, Michahelles F. AppAware: Which mobile applications are hot?. 12th international conference on Human computer interaction with mobile devices and services. Association for Computing Machinery, New York (NY), USA. 2010: 431– 434.

11. Stoyanov SR, Hides L, Kavanagh DJ, Zelenko O, Tjondronegoro D, Mani M. Mobile app rating scale: a new tool for assessing the quality of health mobile apps. JMIR MhealthUhealth. 2015;3(1):e27.

12. Stoyanov SR, Hides L, Kavanagh DJ, Wilson H. Development and Validation of the User Version of the Mobile Application Rating Scale (uMARS). JMIR MhealthUhealth. 2016;4(2):e72.

13. The Mobile App Rating Scale (MARS) and its Application to the Development of a Youth. URL: https://www.c4tbh.org/seminar/title-tba-5/. Assessed September 9, 2020.

14. Gwet KL. Handbook of inter-rater reliability: the definitive guide to measuring the extent of agreement among multiple raters, 3rd edition. 3rd ed. Gaithersburg: Advanced Analytics, LLC; 2012.

15. Menaspà Paolo. Effortless activity tracking with Google Fit. British journal of sports medicine. 2015;49(24):1598.

16. Weber MB, Ranjani H, Staimez LR, et al. The Stepwise Approach to Diabetes Prevention: Results From the D-CLIP Randomized Controlled Trial. Diabetes Care. 2016;39(10):1760–1767.

17. Diabetes Prevention Program (DPP) Research Group. The Diabetes Prevention Program (DPP): description of lifestyle intervention. Diabetes Care. 2002;25:2165–2171.

18. Mor Nachiket. Information Technology for Primary Healthcare in India. 2020. DOI:10.13140/RG.2.2.25513.13925.

19. Mani M, Kavanagh DJ, Hides L, Stoyanov SR. Review and Evaluation of Mindfulness-Based iPhone Apps. JMIR MhealthUhealth. 2015;3(3):e82.

20. Bardus M, van Beurden SB, Smith JR, Abraham C. A review and content analysis of engagement, functionality, aesthetics, information quality, and change techniques in the most popular commercial apps for weight management. Int J BehavNutr Phys Act. 2016;13:35.

21. Arnhold M, Quade M, Kirch W. Mobile applications for diabetics: a systematic review and expert-based usability evaluation considering the special requirements of diabetes patients age 50 years or older. J Med Internet Res. 2014;16(4):e104.

22. Ranjani H, Nitika S, Ranjana RM, Sandhya R, Mohan V, Nalini S. Impact of NCD text messages delivered via an app in preventing and managing lifestyle diseases - results of the ‘myarogya’ worksite-based effectiveness study from India. Journal of Diabetology. 2020;11.

23. Muralidharan S, Ranjani H, Anjana RM, Allender S, Mohan V. Mobile Health Technology in the Prevention and Management of Type 2 Diabetes. Indian J Endocrinol Metab.2017;21:334–340.

24. Shuhan He. mHealth App Development and Standardization of Innovative Technologies. URL: https://conductscience.com/mhealth-app-development-current-practices-and-future-perspectives/. Assessed December 2, 2020.

25. Kazemi A, Salmani H, Shakibafard A, Fatehi F. New and Emerging Mobile Health (mHealth) Technologies and Solutions: A Horizon Scanning Study. Front Health Inform. 2019; 8(1): e17.

26. Stephan LS, Dytz Almeida E, Guimaraes RB, et al. Processes and Recommendations for Creating mHealth Apps for Low-Income Populations. JMIR Mhealth Uhealth. 2017;5(4):e41.

27. L. Nurgalieva, D. O’Callaghan, Gavin Doherty. Security and Privacy of mHealth Applications: A Scoping Review. IEEE Access 2020. PP. 1–1. 10.1109/ACCESS.2020.2999934.

